# Conceptualising care pathways for neglected tropical diseases in sub-Saharan Africa: A systematic scoping review

**DOI:** 10.1101/2024.09.13.24313492

**Authors:** Sandrena Ruth Frischer, Eloise Ockenden, Fabian Reitzug, Michael Parker, Goylette F. Chami

**Affiliations:** Big Data Institute, Nuffield Department of Population Health, University of Oxford, Oxford, UK; Ethox Centre, Nuffield Department of Population Health, University of Oxford, Oxford, UK

## Abstract

**Background:** There is a lack of synthesized evidence on how best to support care continuity for neglected tropical diseases (NTDs) in sub-Saharan Africa (SSA).

**Methods:** To identify all SSA NTD care pathways, we conducted a systematic search of six scientific databases from inception to 18 February 2023. All studies were required to include care pathways for NTDs, defined at minimum as having both NTD diagnosis and treatment or referral to treatment for care continuity. Using an iterative approach to establish an understanding of care pathways relevant for NTDs, concept development drew from data extraction of variables relating to study characteristics; approaches to diagnosis, treatment, and referrals as pathway components; barriers to care and strategies for their resolution; and ethical challenges.

**Findings:** Searches returned 2178 studies where after de-duplication and eligibility screening, 164 were systematically reviewed. Medical referrals were used to support care continuity in 62.2% (102/164) of studies, and community health workers played roles in patient care in 22.6% (37/164) of studies. Only 6.7% (11/164) of studies explicitly mapped care pathways, none of which were for NTDs where preventive chemotherapy is the primary management strategy. The majority of studies (82.9%; 136/164) presented unmapped care pathways, which were primarily biomedical research studies that provided diagnosis and/or treatment of NTD patients (66.9%; 91/136). Of biomedical research studies, 36.3% (33/91) described strategies to support continuity of care. While there is no singular care pathway for all NTDs in SSA, we proposed a conceptual framework relevant for implementation of biomedical research.

**Interpretation:** Further research is needed on morbidity management of NTDs typically addressed through mass drug administration in sub-Saharan Africa. To conduct this research, there is a need for guidance on how research sponsors should collaborate with local health systems to enable continuity of care in low-income, NTD-endemic settings.

**Key messages:** *What is already known on this topic:* Providing continuity of care for neglected tropical diseases (NTDs) in the low-income settings where NTDs are endemic is challenging due to a variety of factors, particularly for chronic NTDs requiring morbidity management.

*What this study adds:* All NTDs require patient care, yet NTDs are unjustifiably divided into morbidity management NTDs and preventive chemotherapy NTDs. Further research on morbidity management is needed on NTDs typically managed through mass drug administration of preventive chemotherapies. We show that implementation of biomedical research on NTDs creates distinct care pathways that connect research participants with local health systems for continuity of care. These pathways provide a conceptual framework to understand care continuity during biomedical research as a practical ethical issue that merits further exploration, and illuminates how biomedical research studies might support care continuity for NTD patients such as through provision of transport, collaboration with community health workers, or structural support to local health system service providers.

*How this study might affect research, practice or policy:* This research opens avenues to create tools to assist researchers in providing care in resource-constrained settings. Researchers should consider care pathways for NTDs as a tool to understand local contexts of health seeking and care delivery prior to research implementation. Through these pathways, researchers can understand potential gaps in care continuity during research implementation, and determine what support may be required to address these gaps and facilitate research activities in low-income settings.

## Introduction

Neglected tropical diseases (NTDs) include more than 20 diseases associated with poverty and prevalent in settings where access to healthcare may be severely constrained (1). NTDs are endemic in many low and middle-income countries (LMICs), with the highest NTD burden globally in sub-Saharan Africa (SSA) (2). For many NTDs, symptoms are initially non-specific and may only become evident upon progression to severe disease (3). Per the the WHO 2023 NTD Roadmap, there is a push from vertical, NTD-specific programming towards integration of NTD care within national health systems (4). To understand the different routes NTD patients may navigate in accessing care, or lack thereof, in resource-constrained LMIC contexts, care pathways have proven a useful framework (5–8). Care pathways and related pathway frameworks, such as patient pathways, outline the journey a patient may take in respect to a specific condition based on patient preferences and decision making, from care seeking and diagnosis to treatment and follow up (9). By emphasizing patient experiences of illness and agency in care seeking preferences, care pathways are well suited for understanding healthcare seeking in real world settings where health systems infrastructure may be limited and recommended care for complex conditions requiring integrated case management may not be available (10).

Using a pathway framework to conceptualise continuity of care, McCollum et al conducted a scoping review on Severe and Stigmatizing Skin NTDs (SSSDs) in SSA to explore barriers to accessible care for NTD patients (11). This review focused on understanding patient experiences accessing the health system to bolster efficient health service delivery. However, this review was limited to select chronic NTDs, and did not include NTDs for which mass drug administration (MDA) of preventive chemotherapy (PC) is the primary control strategy (PC-NTDs). Additionally, there is a need to understand how these barriers are navigated during implementation of biomedical research on NTDs, particularly in the context of health system integration. In this systematic scoping review, we explore all NTDs and concepts of care pathways in SSA to address the following questions: 1) What are the care pathways when managing NTDs in SSA, including during biomedical research implementation? and 2) how is care continuity facilitated within these pathways?

## Methodology

This review followed the Joanna Briggs Institute methodological guidance for conducting systematic scoping reviews, with reporting in line with the 2018 Preferred Reporting Items for Systematic Reviews and Meta Analyses extension for Scoping Reviews (PRISMA-ScR) (S1 Text) (12,13). The full review protocol was published on 28 February 2023 on the Open Science Framework (14).

### Search Strategy

Guided by a 2019 scoping review on patient pathways within integrated healthcare (10), two researchers (SRF and GFC) developed the search string through discussion and iterative search piloting using terminology relating to care pathways. Given multiple understandings and varying applications of “care pathways” terminology and similarity with patient care concepts such as the “care continuum”, “treatment cascade,” and “patient journey”, we opted to include all terminology identified as potentially synonymous or relating to care pathway experiences. We also elected to include “referrals” in the search string to explicitly incorporate how patients move between pathway components, such as from suspect case assessment or diagnosis to follow up. However, terminology for individual components of the pathway such as “diagnosis” and “treatment” were omitted to constrain the search string to the pathway as a whole and its connective processes. All NTDs as recognized by the WHO were included in the search string. This decision allows for comparison of similarities and differences of pathways across NTDs as a group of diverse diseases and symptoms that share the difficulties of political deprioritisation and endemicity in settings of extreme poverty, including challenges accessing diagnosis and treatment. We restricted geographic scope to countries in SSA as categorised by the World Bank to focus on healthcare delivery contexts specific to the region with the highest NTD burden, and to ensure comparisons across similar health systems. The broad search string included “patient pathways” AND “neglected tropical diseases” AND “sub-Saharan Africa”. Using the broad string with adaptations applied as necessary, one researcher (FR) searched the following databases on 16 February 2023 without year restrictions: PubMed/MEDLINE, Embase, Global Health, Global Index, Medicus, and Web of Science Core Collection – Science Citation Index Expanded. The same researcher (FR) also searched the Cochrane Central Register of Controlled Trials (established 1996-present) for randomised controlled trials before uploading all resulting articles into Covidence (Veritas Health Innovation, Melbourne, Australia) for deduplication and eligibility screening. The full string and table of database hits is available in S2 Text.

### Screening

Two researchers (SRF and EO) screened all titles and abstracts resulting from the search. The full text of all eligible studies was then retrieved, where available. All full texts were screened by one researcher (SRF), with a random 10% verified by a second researcher (EO). Disagreements between researchers were resolved through discussion and review of the full text. Where applicable, studies describing the same population at the same time point were selected based on the study providing the most information. Only texts in English or with a published translation were included at the full text stage. Only original research articles with primary data collection involving qualitative, quantitative, or mixed research methodologies were included. Analyses of routine electronic health data were excluded, with no other restrictions in study design. There were no age, sex, or other demographic restrictions in the study population beyond residence in SSA.

All studies were required to include care pathways for NTDs. As there is no standardized definition of what a care pathway entails, we initially defined care pathways as having both NTD diagnosis and treatment or referral to treatment for care continuity as identifiers of care pathways. Diagnosis entailed patient awareness of illness through feedback of clinical findings or self-reporting of symptoms, conditions, or morbidities. Treatment included initiation or follow up at formal, government-regulated providers or informal providers, such as local drug shops and traditional healers. To centre patient perspectives, we required that qualitative studies describing experiences of disease management included study participants with the NTD of focus. Given the paucity of literature on care continuity more broadly for NTDs, we did not require that included studies explicitly state or depict care pathways diagrammatically. Rather, by requiring NTD diagnosis and treatment or referral, we included pathways that were either unmapped given the aforementioned components, or clearly mapped, such as through a diagram or a list of ordered pathway components. Following full text screening, all studies excluded at the full text stage were re-reviewed by SRF to categorise reasons for exclusion.

### Data extraction and concept development

Prior to data extraction, SRF and GFC developed an initial data dictionary and table for extracted variables using Microsoft Excel. We then split data extraction into four stages. In the first stage, we extracted study design and aims, country, NTD of focus, study population. In this stage, we also attempted to derive a singular care pathway from each study for synthesis. However, given the diversity of patient preferences and decision making along with the range of experiences with follow up care and varying study designs, there was no singular pathway that could be derived from all included studies, and we determined this would not be the most appropriate methodology for capturing patient experiences with care continuity. Thus in the second stage, we iterated the extraction methodology to instead capture how care pathways are depicted, themes common to care pathways, and the types of pathways that can emerge in different context. In this second stage, we also began conceptualizing what care pathways can be and what they are not. In the third stage, SRF identified all studies that explicitly mapped care pathways to determine concepts to be explored further. In the fourth and final stage, SRF revised, piloted and iterated the data dictionary and extraction table to capture these concepts before proceeding to extract data from all studies included in this review (15,16). The data dictionary is available in S3 text. The variables we chose to extract in this final stage aligned with following categories: study participants and care providers, including community health workers (CHWs); components of mapped care pathways, including pathway name, depiction, and starting and ending points; components of unmapped care pathways, including approaches to diagnosis, treatment, and referrals; continuity of care, including barriers to care and strategies or recommendations for their resolution; and ethical challenges identified by study authors. One researcher (SRF) extracted full texts in all stages. At the end of the fourth stage, a second researcher (EO) verified extraction of a random 10% of included studies, resulting in the final extraction table (S1 Dataset). Additionally, while we began with “patient pathways” as the primary terminology, during this concept development we shifted to “care pathways” as a more representative term that can encompass multiple perspectives beyond the patient.

### Analysis

Summary statistics for study characteristics are presented in line with guidelines for systematic reviews without meta-analysis (SWiM)(17). All studies were categorized based on study aims and how the care pathway was represented. For studies describing patient experiences as participants in biomedical research, we employed a “best fit” framework synthesis to outline an example pathway applicable to this context (18). To explore differences between NTDs across study aims and pathway representations, NTDs were grouped into three categories based on WHO care management approaches and treatment contexts: PC-NTDs (schistosomiasis, trachoma, scabies, and lymphatic filariasis), NTDs which cannot be treated through MDA of PC and require morbidity management (leprosy, buruli ulcer, podoconiosis, and human African trypanosomiasis (HAT)) and NTDs caused by animal bite injury (rabies and snakebite) (2,23). Within each categorization of study aim and care management approach, thematic synthesis was used to code the data and develop descriptive themes (19).

### Patient and public involvement

There was no patient or public involvement in the methodology of this review.

### Ethical approval

As this study reviewed existing published literature, no ethical approval was required.

## Results

Figure 1 details the search process for this systematic scoping review. The initial database search resulted in 2178 studies. Following deduplication, 1257 titles and abstracts were screened and 963 were deemed ineligible, leaving 294 studies. After full text retrieval and screening, 164 studies were available and met the inclusion criteria (S4 Text).

**Figure 1:**
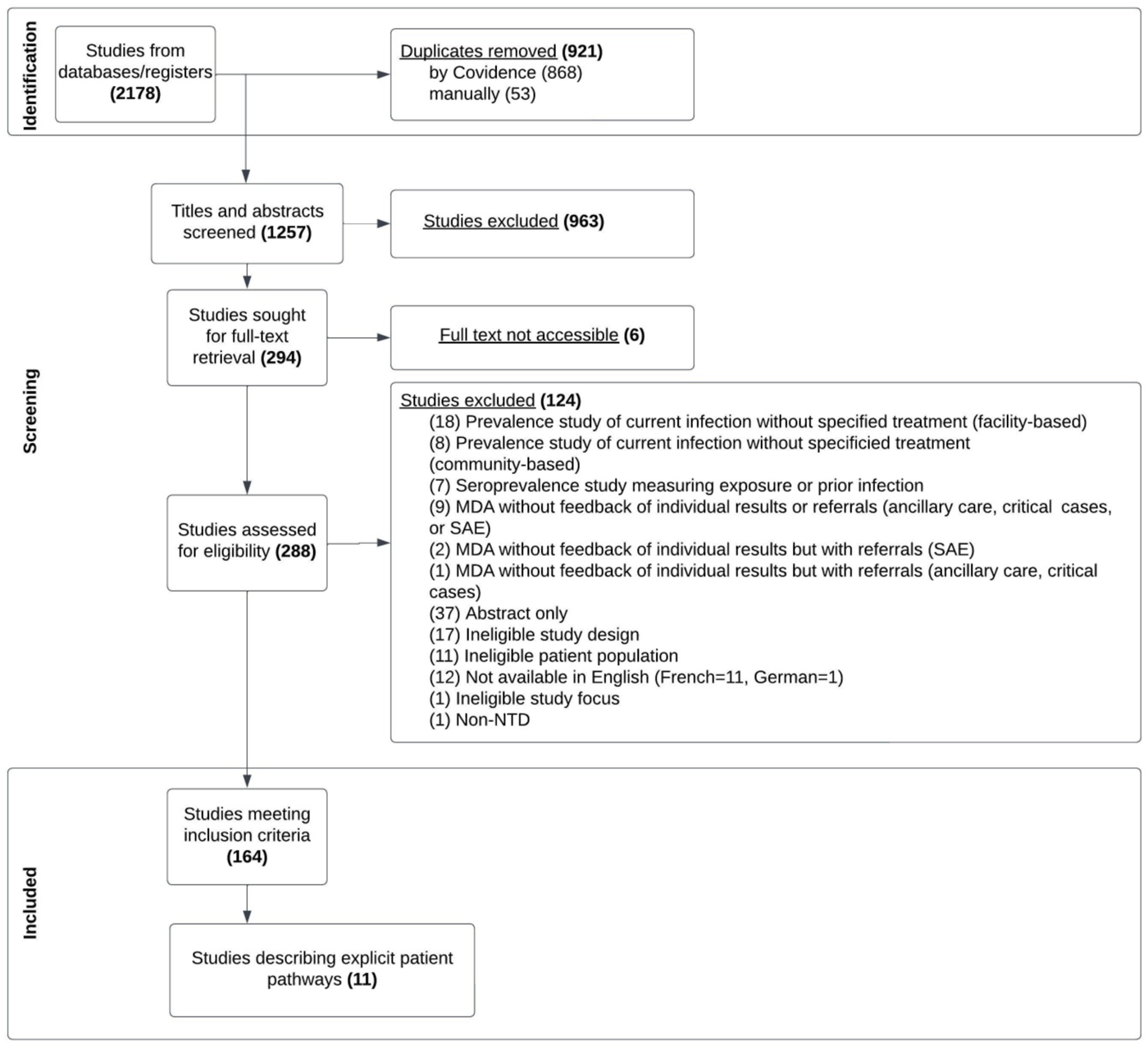
PRISMA Study Selection Flowchart. The screening and review process is split into three stages: 1) the identification of articles through an initial search, 2) the screening of articles following predefined inclusion and exclusion criteria and 3) the final included articles for data extraction. MDA: Mass Drug Administration, SAE: Severe Adverse Events.

All extracted variables from 164 included studies are available in S1 Dataset. Figure 2 describes the geographical distribution in SSA of all included studies, with 62.5% (30/48) of SSA countries represented. The most common study countries were Ethiopia (17.6%; 29/164), Ghana (12.2%; 20/164) and Nigeria (11.6%; 19/164), with 2% (4/164) studies taking place in regions extending across international borders (Uganda and Kenya; Niger, Nigeria, Cameroon, and Chad).

**Figure 2:**
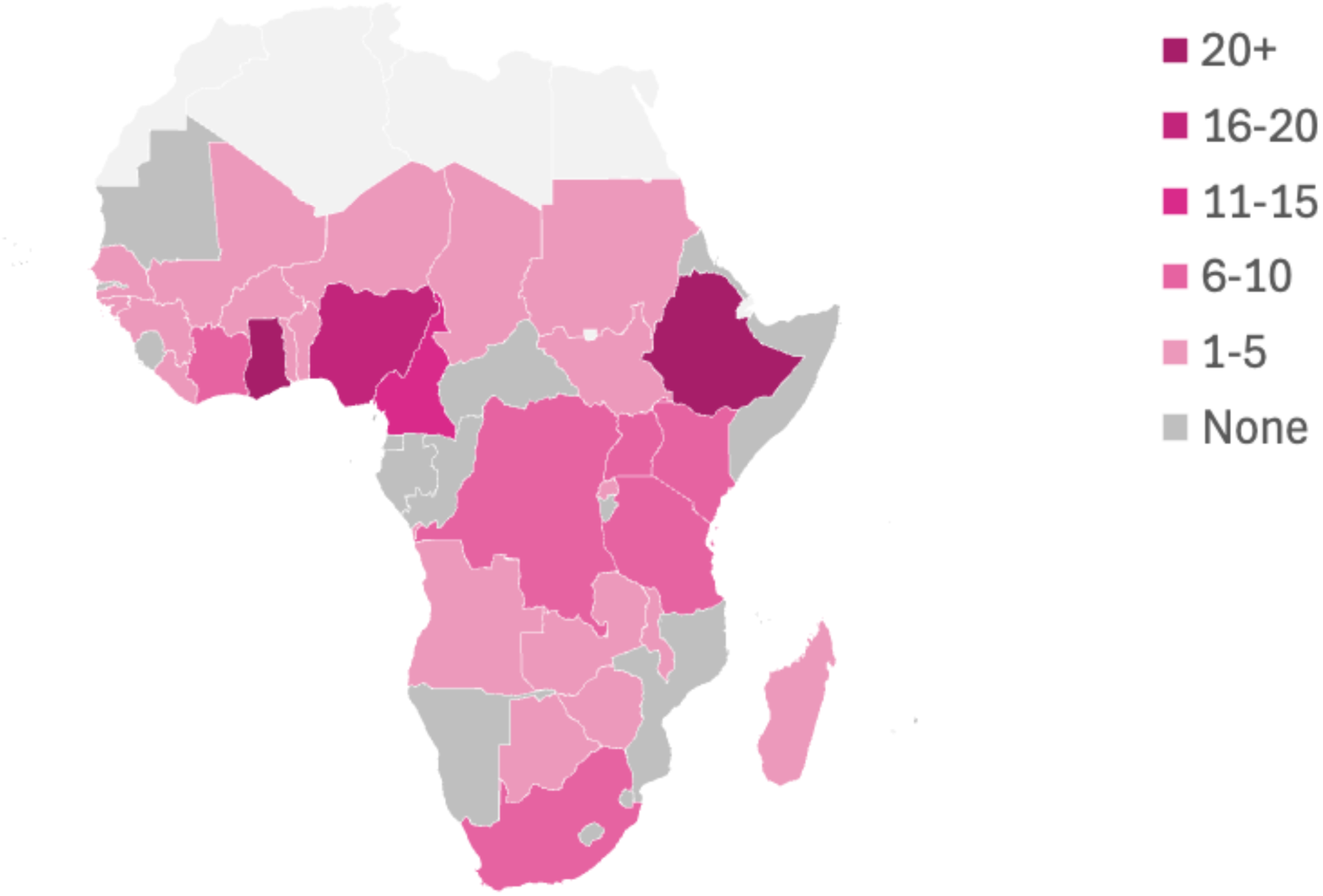
Geographic distribution of studies in sub-Saharan Africa.

Study publication years ranged from 1970 to 2023. More than 71% (117/164) of included studies were published from 2012, the year of the London declaration to control or eliminate at least 10 NTDs by 2020 (20). Total number of study participants varied substantially with study design, with qualitative studies describing in-depth NTD patient experiences, averaging 57 (SD=85) participants. Non-qualitative, non-case report studies averaged 8283 (SD=32,016) participants, typically as participants in mass screening campaigns. **Table 1** summarises key study characteristics.

**Table 1:**
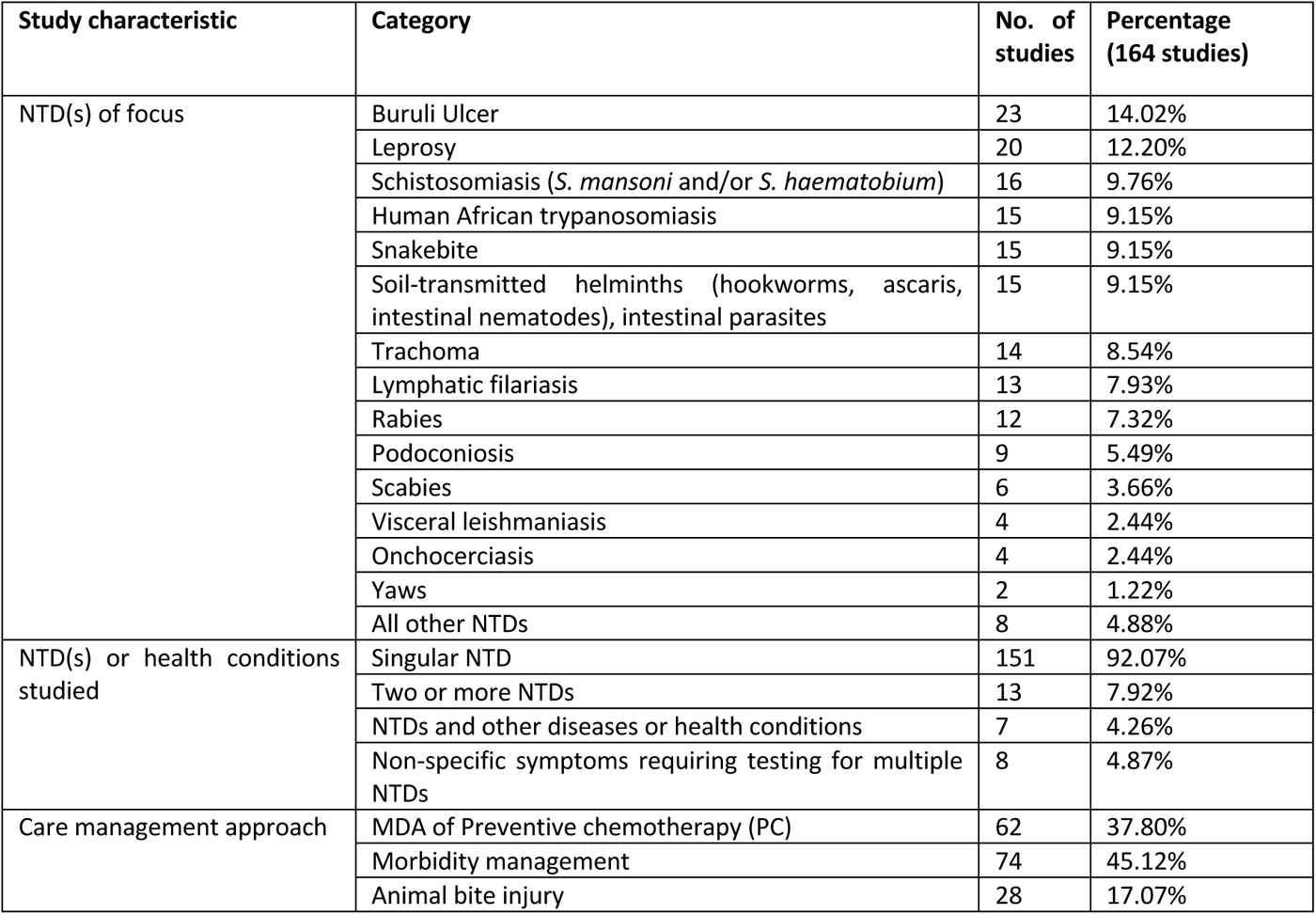
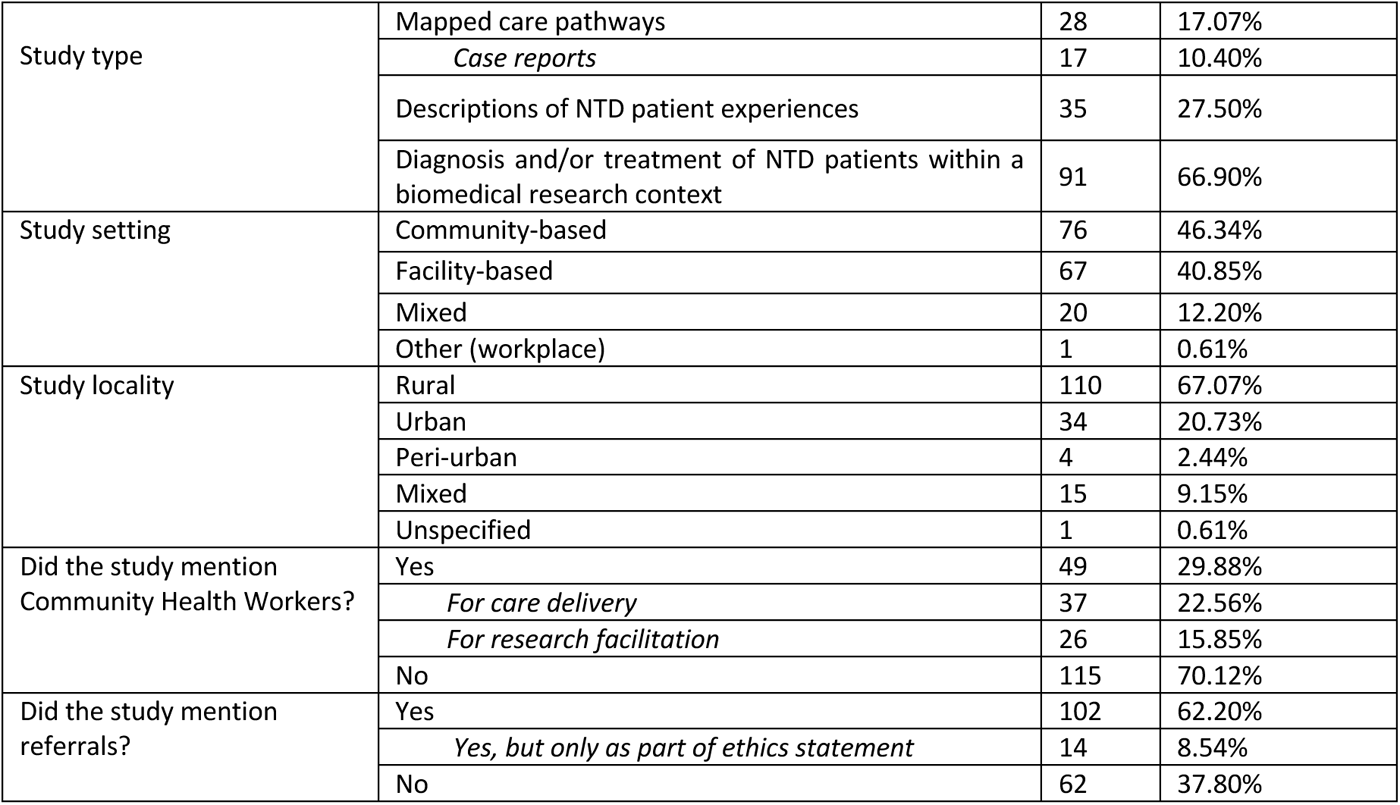
NTD distribution and study characteristics summary. The total number of all included studies was 164. Care management approach reflects categorization and primary treatment regime as defined by the WHO (2,23). All other NTDs include one study each for chikungunya, Guinea worm, cystic echinococcosis, cutaneous leishmaniasis, dengue, mycetoma, and neurocysticercosis. There were 7/164 (4.26%) looking at NTDs alongside comorbidities or other non-NTD health conditions (malaria (2), HIV (2), diabetes (1), pregnancy (1), intimate partner violence (1)). There were also 8/164 (4.87%) studies which targeted non-specific symptoms where NTDs were discussed as a diagnosis (anaemia (3), eye disease or blindness (2), lymphoedema (1), common skin diseases (1), and central nervous system infections (1)).

### Community health workers

Nearly one third of included studies (49/164) mentioned involvement of community health workers. Among these 49 studies, there was substantial variation between CHW terminology, strategies for identification, and work expectations. CHWs were identified by 28 different terms, with 38.8% (19/49) studies using the term community health workers (CHWs), 8.2% (4/49) using community health volunteers (CHVs), 6.1% (3/49) using health extension workers, and 6.1% (3/49) using community drug distributors. Only 28.6% (14/49) of studies with CHWs provided CHW definitions or selection strategies. Among these, 28.6% (4/14) were elected based on community support, 28.6% (4/14) were repurposed from other activities, such as implementation of MDA, and 42.9% (6/14) were required to have some form of healthcare experience, such as being a retired health professional. CHW responsibilities encompassed both roles that CHWs played in care delivery and in research facilitation. CHWs played roles in patient care in 22.6% (37/164) of studies overall, and in studies 75.5% (37/49) of studies mentioning CHWs.CHW responsibilities for patient care included health promotion, support with daily self-treatment for NTDs requiring morbidity management, active case finding and case identification, and provision of medical referrals. In the 53.1% (26/49) of studies where CHWs were described as providing roles in research facilitation, typical responsibilities involved community engagement and participant enrolment, including taking of informed consent (21–31), participant recruitment through either enumeration of households or registration of presumptive cases (32–35), and interpretation support to communicate with study participants in local dialects (36,37). In the 55.1% (27/49) of studies describing training for CHWs to perform their roles, training ranged from care strategies including early stage NTD case identification (25,26,38–42), to basic treatment provision (41,43), to capacity building for research facilitation through training in survey procedures and interviewing techniques (28,29,36,44).

### Medical referrals

Medical referrals were described in 62.2% (102/164) of included studies. Referrals were less described in studies on PC-NTDs (38.2%; 39/102) than studies on NTDs requiring morbidity management or resulting from bite injury (61.7%; 63/102). Reasons for referral included further assessment and confirmatory diagnosis in 20.5% (21/102), treatment of any sort in 53.9% (55/102), speciality care for management of complex cases in 35.3% (36/102), and treatment requiring medication that was not available locally in 10.8% (11/102). While studies generally did not report whether medical referrals resulted in further medical care, two studies described incomplete referrals due to patient deaths during transit. In several studies, provision of medical referrals was problematized given limitations in healthcare accessibility. For example, one evaluation of a complex intervention on buruli ulcer control in Ghana sought to facilitate early disease recognition and referral (45). For participants in this study, an increase in referrals and a decrease in treatment non-completion was associated with increasing healthcare accessibility and supporting the availability of medical commodities at the treating health facility, engaging with providers to ensure acceptable treatment of study participants, provision of transport for accessible care, and connecting patients with free and affordable health services.

### Care pathway representation and study categorisation

Given that there was no singular pathway that could be derived from all included studies, studies were categorised based on whether care pathways were either explicitly mapped or unmapped. Studies determined to have mapped care pathways described the varying routes NTD patients may go through in accessing or attempting to access care from patient perspectives. Mapped pathways also provided terminology to identify the care pathway as a concept in itself beyond the simple inclusion of diagnosis and treatment or referral as pathway components, such as “health seeking pathway” or “illness narrative.” In identifying mapped pathways, we recognized several distinctions between mapped care pathways and diagnostic or treatment algorithms. Studies describing algorithms involved sequences of diagnostic steps to arrive at a confirmatory diagnosis. Conversely, mapped care pathways highlighted actual patient experiences along the care continuum, which deviated from clinically recommended diagnostic steps. While clinical case reports meet this criteria, we opted not to include these publications as examples of mapped pathways and analyse them separately given the facility-based context and distinct study design. Unmapped pathways were defined as meeting the care pathway criteria of including NTD diagnosis and referral, but without an explicit connection between pathway components represented through a diagram, list, or narrative.

Among unmapped care pathways, studies were also categorised based on if the study either 1) described the lived experiences of NTD patients, or 2) provided diagnosis and/or treatment of NTD patients and therefore an element of patient care within a biomedical research context. Studies describing NTD patient experiences were typically cross-sectional and used mixed methodologies or qualitative data collected through interviews or focus groups to describe the substantial barriers NTD patients face when attempting to access care and follow up. Biomedical research studies that provided an element of patient care included the following study designs: cross-sectional prevalence studies, randomised controlled trials (RCTs), case-control studies, or cohort studies, evaluations of complex interventions. Within each of these categories, NTDs were described by care management approach.

#### Mapped care pathways

We identified mapped care pathways that were not case studies in 6.7% (11/164) of included studies. Summary characteristics of these mapped pathways are presented in **Table 2**. Mapped pathways were represented in a range of visual and geographic formats including flowcharts, narratives, and maps, with varying starting points ranging from exposure, infection, onset of symptoms, to care seeking. Of all 11 mapped pathways, 63.6% (7/11) were for NTDs requiring morbidity management and 36.4% (4/11) NTDs caused by animal bite injury. No mapped pathways were identified for PC-NTDS. All seven studies for NTDs requiring morbidity management referenced misdiagnosis as a key pathway component. HAT was the disease of focus in 27.2% (3/11) of mapped pathways, where key barriers to care continuity in the pathway included difficulty of diagnostic testing given non-specific symptoms, lack of testing availability, and false positives (5–7). SSSD NTDs were the focus of 36.4% (4/11) studies, all of which including exploration of patient experiences with chronic buruli ulcer (8,36,46,47). One study mapped SSSD NTD care pathways in rural Liberia using narratives of patients living with buruli ulcer, leprosy, lymphatic filariasis, or onchocerciasis to examine the protracted trauma of living with a disabling, stigmatizing condition with no straightforward treatment in a resource-constrained setting (36). The 36.4% (4/11) studies on animal bite injury all referenced the need for prompt biomedical response despite limited availability of treatment (21,22,48,49). There was substantial variation among mapped pathway endings, from arrival at the health facility, to diagnostic tests conducted, to diagnosis confirmed, to start or end of treatment, to lack of ending given perpetuation of chronic disease. Only 18.2% (2/11) of these studies continued the pathway beyond the start of treatment to reflect treatment adherence and care continuity.

**Table 2:**
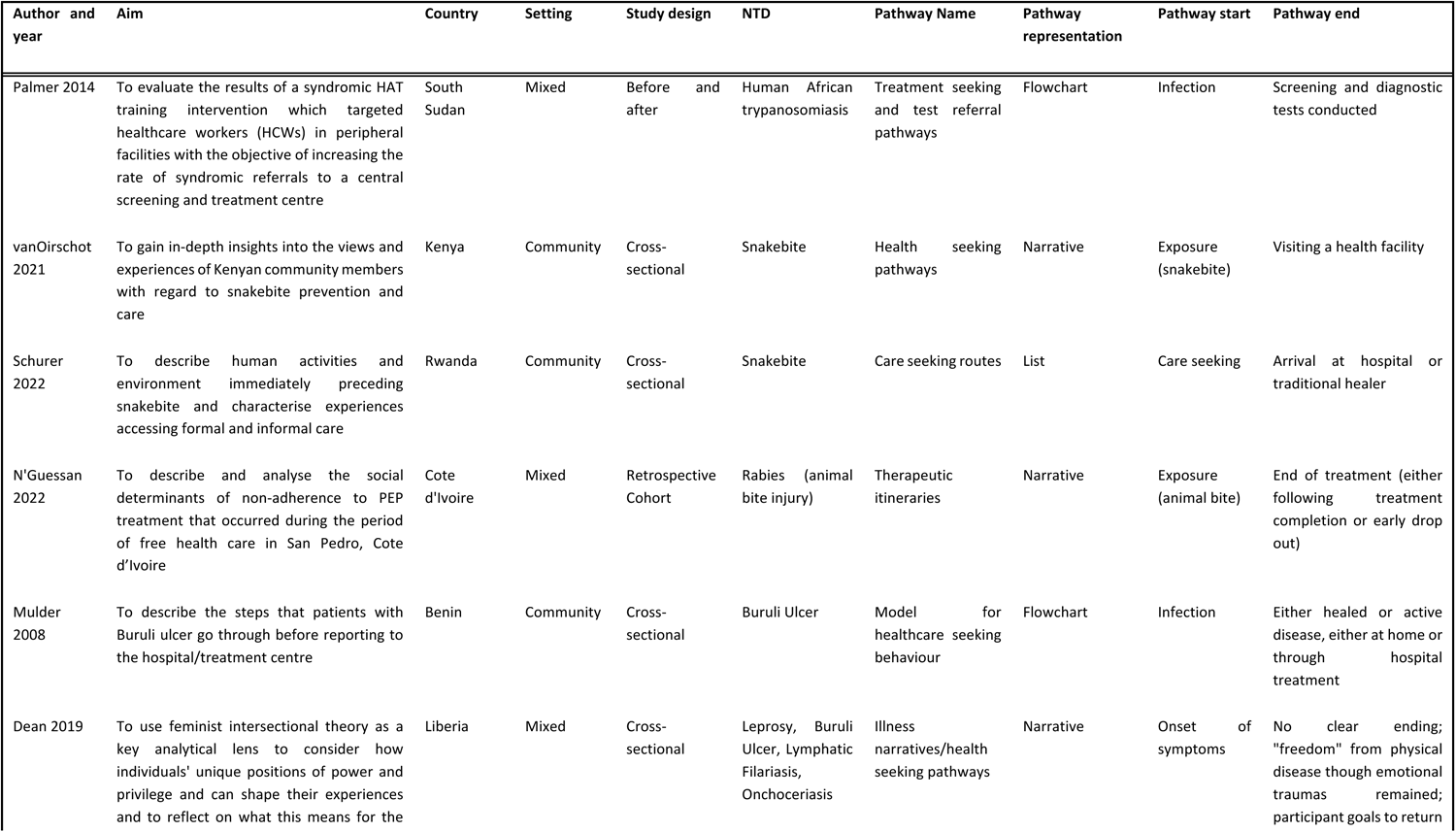
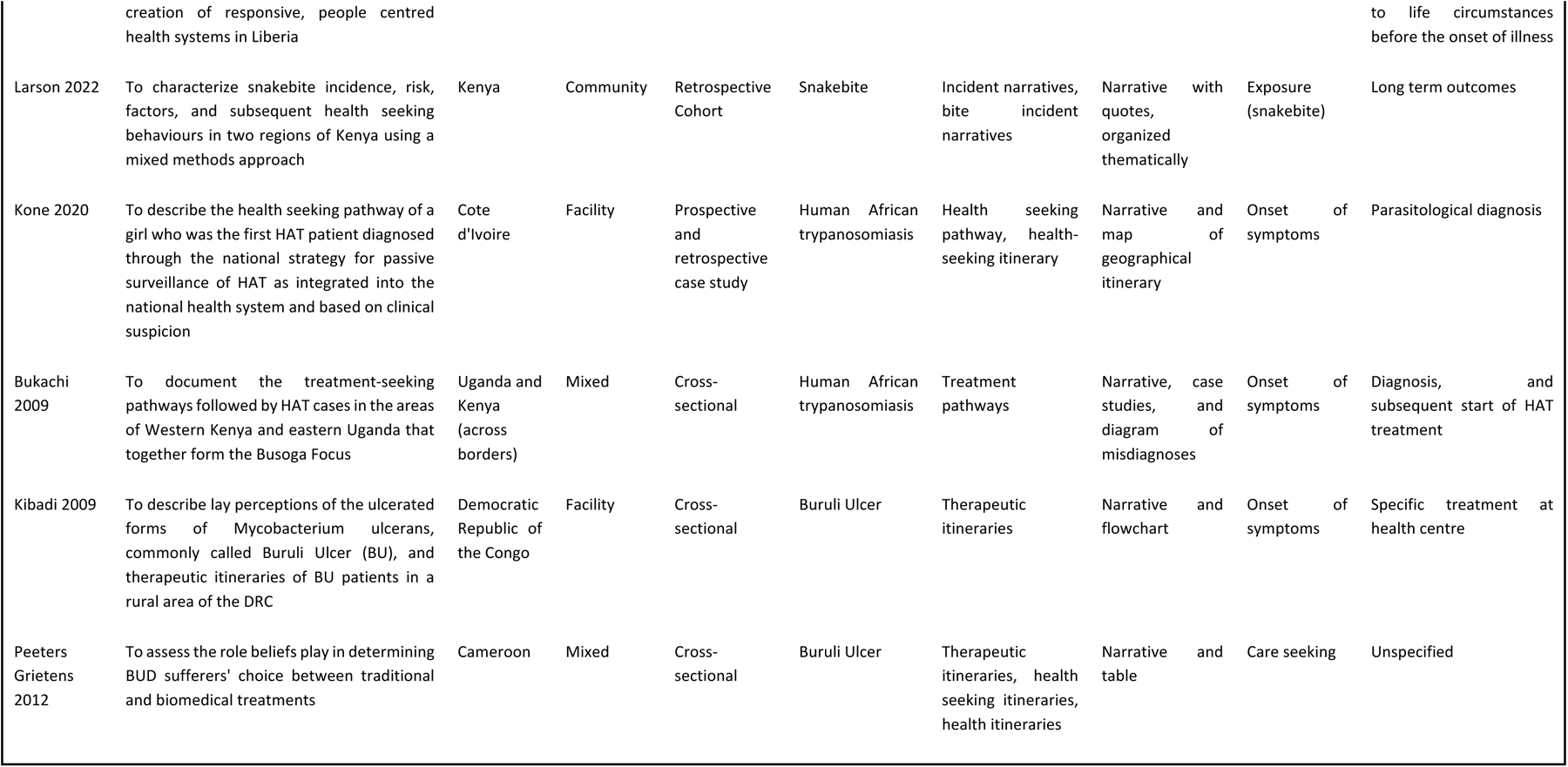
Study characteristic summary table for studies describing mapped care pathways.

Among included texts, 10.4% (17/164) were clinical case reports. All clinical case reports were facility-based and provided accounts of clinical management for 1-2 individuals from care seeking through diagnosis and treatment from clinician perspectives. Only 18% (3/17) of case reports focused on PC-NTDs, all of which were on schistosomiasis. All case reports provided insight as to the care patients received upon arrival at the health facility. Rarely was this a linear path, as patients presenting with non-specific symptoms frequently required several rounds of diagnostic testing and trial treatments. Given the nature of clinical case reports as a medium for describing clinical decision processes and patient outcomes in facility settings, the conditions represented often described complex or atypical cases. For example, in the three case reports on patients with schistosomiasis, two case reports described cancer diagnoses associated with chronic schistosomiasis infection (50,51). Several case reports supplemented accounts of clinical care with exploration into the epidemiological context preceding diagnosis and suspected barriers to care. One such case report on HAT combined description of case management with a retrospective epidemiological investigation to understand the manifold barriers the patient had faced prior to accessing care at the present facility (6).

#### Unmapped care pathways

The majority of included studies (82.9%; 136/164) presented unmapped care pathways. Within these, 35 studies (25.7%) described patient experiences of diagnosis and treatment. These studies referenced financial, geographic, social, and structural barriers to care continuity alongside non-linear care processes, where patients often sought symptom management prior to receiving a diagnosis, and obtaining a diagnosis did not necessarily result in treatment. Of these descriptive studies, 62.8% (22/35) were on NTDs requiring morbidity management, while NTDs resulting from animal bite injury and PC-NTDs comprised 11.4% (4/35) of these studies. PC-NTDs were the focus of the remaining 25.7% (9/35) studies. Of the nine studies on PC-NTDs, 50% (5/10) were on lymphatic filariasis, 40% (4/10) were on schistosomiasis, and one study was on trachoma. Among these 10, five explored barriers to care for treating chronic conditions (32,52–55), two of which described the challenges of care seeking given presentation of non-specific symptoms associated with schistosomiasis infection (54,55). Two studies highlighted the need for and accessibility of surgical services for severe disease for trachoma and lymphatic filariasis, respectively (56,57).

The majority of unmapped care pathways (66%; 91/136) were biomedical research studies that provided diagnosis and/or treatment of NTD patients and therefore an element of patient care. Of these, 52.7% (48/91) of studies were on PC-NTDs, with 81.3% (39/48) of these using cross sectional designs to determine PC-NTD prevalence. The remaining 39.6% (36/91) and 7.7% (7/91) of biomedical research studies focused on NTDs requiring morbidity management and NTDs resulting from animal bite injury, respectively. All care pathways for participants in biomedical research were implicit, i.e. not described diagrammatically.

##### Proposed pathway framework

To map the interconnected components of care pathways for cross-sectional prevalence studies as the most frequent study design, a proposed care pathway for participants in biomedical research is depicted in Figure 3.

**Figure 3:**
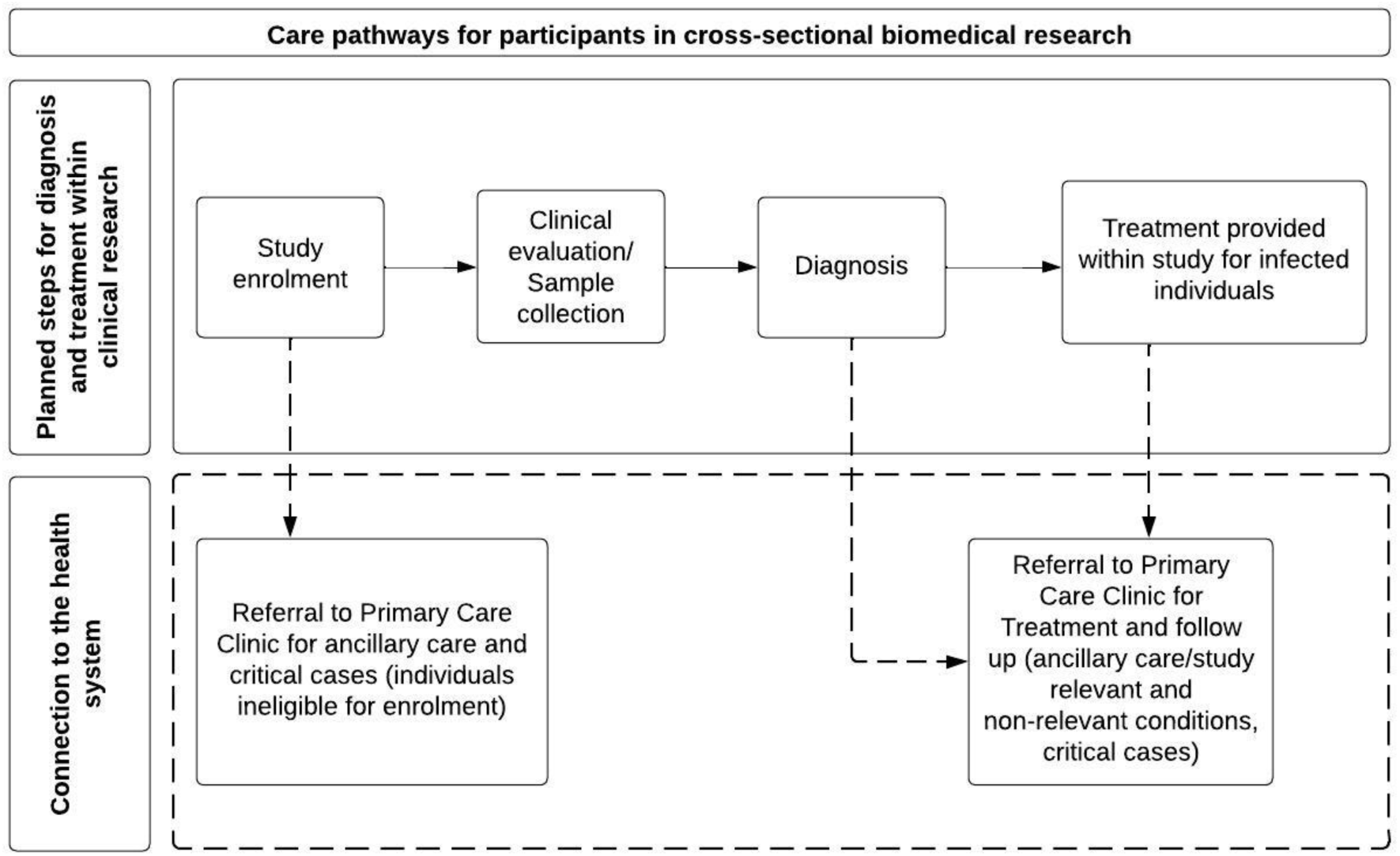
Proposed care pathway for participants in biomedical research.

In Figure 3, the care pathway is defined from the research study perspective and denoted through solid arrows, while dashed arrows indicate when research participants may receive medical referrals to the health system for continuity of care. Through this schematic, care pathways during biomedical research studies can be understood as diagnostic or treatment algorithms, with the critical addition of the complex, non-linear experiences a patient might have while trying to access care through the health system beyond the research context.

##### Ethical challenges and strategies to support care continuity during biomedical research

For participants in biomedical research, only 7.8% (7/91) studies explicitly mentioned any ethical challenges. These included improvements in health facility reputation resulting in a challenging increase in patient load (25), and the need to connect research participants with immediate local support in the event of acute mental distress (58). Additionally, 15.4% (14/91) of studies mentioned treatment or referral for study participants with a positive NTD diagnosis as part of the research ethics statement, and of these, only 14.3% (2/14) described any strategies for follow up. Of all 91 biomedical research studies, 36.3% (33/91) of studies described strategies to support care continuity and follow up of NTD patients. Strategies included provision of food to patients or transportation to access treatment (45,59–61), training for CHWs and support from CHWs in delivering community-based treatment (23,38,39,44,62,63), use of additional and alternative modes of communication, such as SMS (64), and adjustments to accessibility of treatment and speciality care, such as provision of free surgical services or use of mobile clinics to make care more accessible in community settings (40,65–69). Strategies were also described for building relationships between research studies and local clinics (24–26,38–41). In one study in Nigeria, clinics to receive referrals from study participants were visited every five weeks by research project staff to discuss preparedness to receive cases (24). In several complex interventions, strategies for care continuity extended beyond study participants to address structural or health systems factors. One study on morbidity management for lymphatic filariasis in Zambia took such an approach, highlighting how study funds were directed to improving the health facilities to receive referrals for care continuity (53). In this study, these investments were framed as elements of health system strengthening with potential to improve care delivery within and beyond research implementation.

## Discussion

Providing care for NTDs requires engaging with resource-constrained health systems in the low-income settings where NTDs are endemic, such as in many parts of SSA. Through this systematic scoping review, we conceptualised care pathways for NTDs in SSA by identifying 164 studies that included pathway components of NTD diagnosis and treatment or referral to treatment for continuity of care. The conceptualisations presented in this review suggest that there is no singular care pathway for all NTDs in SSA. Care pathways for NTDs in SSA are marked by complex, nonlinear care seeking experiences, and there is a need to recognize the different care pathways that emerge in different contexts, such as during implementation of biomedical research. Importantly, conceptualisation of care pathways within biomedical research on NTDs in SSA, to our knowledge, has not been done prior to this review. This conceptualisation presents an opportunity to understand the distinct care pathways created during biomedical research that connect research participants with health systems for continuity of care, in contrast with care pathways based in routine health service delivery. Using a care pathway framework, researchers can understand potential gaps in care continuity during research implementation, and determine what support may be required to address these gaps and facilitate research activities in low-income settings.

The financial, geographic, social, and structural barriers to care continuity highlighted in this review broadly align with those identified in the McCollum review on SSSDs (11). This congruence suggests that, given the context of extreme poverty in which NTDs occur, challenges accessing care for SSSDs are shared across all NTDs, particularly for NTDs with chronic and/or non-specific manifestations. The most critical aspect of NTD care may not be in the pre-existing NTD management sub-categorizations, but rather the shared context of neglect and constrained health system capacity. Despite this, this review highlights a relative lack of research on care pathways for PC-NTDs and patient experiences of care continuity for PC-NTDs, compared to NTDs that require morbidity management per WHO guidance. This is most salient in the 11 mapped pathways, none of which focused on PC-NTDs. The limited number of case reports for PC-NTDs stresses this point, given the value of these reports as a source for development of case management strategies. This limited evidence would at face-value appear to suggest that complex care seeking processes and the need for morbidity management are not applicable to PC-NTDs. Yet, as emphasized in the 13 descriptive studies and case reports for PC-NTDs, patients suffering from PC-NTDs do require morbidity management and have complex care seeking experiences that can involve misdiagnosis and loss to follow up. This research gap reflects the limitations of MDA of PC as the predominant management strategy for PC-NTDs, particularly given imperfect MDA implementation that excludes high-risk populations alongside lack of PC-NTD treatment integration within health systems (70–72). For example, chronic schistosomiasis infection due to missed PC administration in schistosomiasis-endemic areas can result in nonspecific gastrointestinal symptoms which may develop into severe gut and liver morbidities, including periportal fibrosis that requires advanced care and follow up (73). However, despite the substantial burden of chronic schistosomiasis infection among impoverished individuals in rural sub-Saharan Africa, there are no clinical care or morbidity guidelines for schistosomiasis (74). To develop care guidelines that extend beyond MDA and include morbidity management, there is a need for further case management research on schistosomiasis and other PC-NTDs.

During research activities, research sponsors have an ethical obligation to make a reasonable effort to meet the health needs of research participants (75–77). This obligation raises complex ethical questions regarding continuity of care in NTD endemic settings of extreme poverty, as the necessary care might not be accessible. Despite this, only 7.8% of biomedical research studies included in this review explicitly mentioned ethical issues of any nature. To bolster discourse on practical ethical issues during biomedical research implementation, the conceptualisation of care pathways presented in this review provides a framework to understand care continuity as a practical ethical issue that merits further exploration. In research-based care pathways, the research environment creates a unique opportunity for NTD diagnosis which might not otherwise be available in a low-income context. However, through referrals to the local health system for care delivery, biomedical research studies may create ethical tensions by exposing participants to challenges for accessing follow-up care while not necessarily presenting solutions. This tension is reflected in prior social research embedded within biomedical research in SSA showing participants referred to the health system for further treatment often are not able to access care through referrals alone (78–80).

To address ethical tensions and better facilitate continuity of care during research activities, this review identifies strategies for provision of additional support from research sponsors to research participants. These strategies include collaborations with CHWs or relationship building between research studies and local clinics to ensure referral preparedness. In some cases, these strategies involve structural support to health system service providers, which can be understood as collateral research benefits to communities where research is taking place (81). Yet, there remain ethical questions regarding what support should be provided to make follow-up care accessible during, and following, biomedical research activities. Open questions include who should provide this support, the type and duration of support, and the resulting implications of this support on the local health system. For example, if a research participant requires care for which the necessary treatment is not accessible, what role should the research sponsor play in facilitating access to care? Should the research sponsor focus on the immediate needs of the research participant, or is there an obligation to address gaps more sustainably by strengthening the broader health system? If the necessary care does not yet exist or has not yet been developed, should the research sponsor play a role in this development? As is true for many NTDs, challenges relating to care continuity for research on conditions with non-specific symptoms become even more relevant when considering multiple potential diagnoses and ancillary care needs that may extend beyond the immediate research scope (80,82). Alongside more established ethical issues during biomedical research in LMICs, such as selection of research participants and discrepancies surrounding standards of care, conducting biomedical research in the low-income contexts where NTDs are endemic necessitates consideration of these practical ethical issues relating to care continuity.

Addressing care continuity during biomedical research on NTDs in low-income settings may have important practical ethical dimensions for CHWs as research stakeholders. As indicated in this review, CHWs often play a role in NTD care pathways and can support facilitation of referrals that connect NTD patients with further care. This representation of CHWs reflects the historical engagement of CHWs in successful NTD program implementation and care delivery in low-income settings (83), and suggests a continued need to strengthen CHW roles within health systems for NTD care delivery. However, this review also highlights how CHWs are engaged in biomedical research implementation in NTD endemic settings of extreme poverty. By leading community engagement activities, or identifying and enroling study participants, CHWs can act as liasons between research participants, communities and sponsors. In these roles, CHWs who support research participants in accessing treatment may end up navigating the aforementioned ethical tensions relating to care continuity, such as limitations in study provision of ancillary care, or lack of means to connect research participants with accessible government healthcare services (84,85). Importantly, this review identified substantial study variation in CHW roles, responsibilities, training, recruitment, and approaches to compensation. This variation reflects the diversity of CHW models across health systems in SSA, alongside a lack of guidance for how CHWs can or should best be engaged to support implementation of biomedical research. To facilitate appropriate and effective research collaborations involving CHWs or other local healthcare providers across distinct, NTD endemic research environments in SSA, this variation highlights the need for understanding care pathways related to the research focus at the site of research implementation, prior to biomedical research activities. This understanding includes determining existing structures for community health service provision, local capacity strengths or needs, and the roles of local stakeholders in care delivery.

### Limitations

As a scoping review, this study did not assess evidence quality or risk of bias in included articles. While this review seeks to capture key components of NTD care pathways in SSA, describing care pathways was not the main objective in the majority of studies. It is possible that several study components extracted for analysis, such as use of referrals and ethical challenges, were simply not reported rather than not done, which may lead to some underreporting in this review. Providing this detail in research publications would aid future researchers in identifying and addressing these issues during research activities. Given the fluid nature of study terminology relating to diagnosis, treatment, and care pathways, the findings of this scoping review may not be entirely exhaustive. Additionally, the English language restriction may have excluded publications from francophone and lusophone SSA or reports in local languages, leading to underrepresentation of care pathways from particular countries.

## Conclusion

The future of NTD care relies on health system integration (4). Yet, practical questions remain on how best to integrate biomedical research studies on NTDs into health systems. In NTD endemic settings of extreme poverty, implementation of biomedical research creates distinct care pathways that connect research participants with local health systems for care. The findings from this comprehensive review inform what considerations may be required to facilitate continuity of care during research on NTDs. To facilitate necessary further research on morbidity management of PC-NTDs and their chronic presentations, there is a need for bioethics research on how research sponsors and local health systems should collaborate to enable continuity of care in SSA and low-income settings more broadly.

## Supporting information

S1 PRISMA SCr Checklist

S2 Search Strings and Databases

S3 Data dictionary

S4 References for included studies

## Data Availability

This systematic scoping review extracted data from openly available published literature. The extracted dataset is available from the authors.

## Acknowledgements

We thank Nia Roberts at the Bodleian Libraries for her help developing the search strategy during the initial stages of the review. We also thank the Chami Group, the Ethox Centre, collaboraters in the Department of Ethics, Law, and Humanities at the Amsterdam UMC and attendees at the 2023 Oxford Global Health and Bioethics conference for their feedback, support, and general advice.

## Declarations

### Author contributions

Conceptualization: **SRF and GFC.** Resources, and supervision: **GFC and MP.** Data curation and validation: **SRF and EO**. Formal analysis, investigation, methodology, validation, visualization, and writing – original draft: **SRF**. Writing – review and editing: **SRF, EO, FR, MP,** and **GFC**. Funding acquisition: **SRF and GFC**.

