## Supplementary material for "Conceptualising care pathways for neglected tropical diseases in sub-Saharan Africa: A systematic scoping review": S2 Search Strings and Databases

**S2 text: Full search string and tables of database hits**

The logic of the search string is as follows:

| **#** | **Search terms** |
| --- | --- |
| **1** | “Patient pathways” |
| **2** | “Neglected Tropical Diseases” |
| **3** | “sub-Saharan Africa” |
| **4** | 1 and 2 and 3 |

This generic search string is adapted to suit constraints and functionalities of different databases. Medline, Embase and Global Health are searched through Ovid (https://www.wolterskluwer.com/en/solutions/ovid). Web of Science is accessed through Clavirate (https://clarivate.com). The exact search string used in each database and interpretations of MeSH terms are provided below.

*Searches and search results (run 2023-02-16):*

**Medline (Ovid MEDLINE®) 1946 to present**

| **#** | **Search terms** | **Results** |
| --- | --- | --- |
| **1** | ("referral" OR "patient pathway" OR "patient-pathway" OR "clinical pathway" OR "care continuum" OR "continuum of care" OR "continuum of patient care" OR "care continuity" OR "continuity of care" OR "continuity of patient care" OR "patient journey" OR "treatment cascade" OR "care cascade" OR "case management" OR "care management" OR "care pathway" OR "treatment pathway" OR "care delivery" OR "care network" OR "link to care" OR "linkage to care" OR "links to care" OR "health seeking" OR "health access" OR "healthcare access" OR "health care access" OR "medical access" OR "care seeking" OR "care access" OR "access to care" OR "access to healthcare" OR "access to health care").mp. | 302603 |
| **2** | ("neglected tropical disease" OR "NTD" OR "schisto*" OR "bilharzia" OR "snail fever" OR "mansoni" OR "japonicum" OR "mekongi" OR "guineensis" OR "s. intercalatum" OR "haematobium" OR "leprosy" OR "hansen’s disease" OR "buruli ulcer" OR "snakebite" OR "mycobacterium ulcerans" OR "chagas" OR "trypanosome cruzi" OR "american trypanosomiasis" OR "dengue" OR "chikungunya" OR "dracuncul*" OR "guinea-worm" OR "guineaworm" OR "echinococc*" OR "foodborne trematodiases" OR "foodborne trematode infection" OR "clonorchiasis" OR "opisthorchiasis" OR "fascioliasis" OR "paragonimiasis" OR "human african trypanosomiasis" OR "sleeping sickness" OR "HAT" OR "trypanosome" OR "leishmaniasis" OR "kala azar" OR "lymphatic filariasis" OR "elephantiasis" OR "podoconisis" OR "wuchereria bancrofti" OR "brugia" OR "onchocerc*" OR "river blindness" OR "rabies" OR "scabies" OR "soil transmitted helminthiasis" OR "roundworm" OR "ascaris" OR "whipworm" OR "trichuris" OR "hookworm" OR "necator americanus" OR "ancylostoma duodenale" OR "taenia*" OR "cysticercosis" OR "tapeworm" OR "mycetoma" OR "chromoblastomycosis" OR "deep mycoses" OR "trachoma" OR "endemic treponematoses" OR "treponem*" OR "yaws" OR "framboesia" OR "endemic syphilis" OR "bejel" OR "pinta").mp. | 295372 |
| **3** | (Africa OR "Sub-Saharan Africa" OR Angola OR Benin OR Botswana OR "Burkina Faso" OR "Burkina Fasso" OR "Burkina Faso" OR "Upper Volta" OR Burundi OR Urundi OR Cameroon OR Cameroons OR Cameron OR Camerons OR "Cape Verde" OR "Cabo Verde" OR "Central African Republic" OR Chad OR Comoros OR "Comoro Islands" OR Comores OR Congo OR Zaire OR "Cote d'Ivoire" OR "Ivory Coast" OR "DRC" OR "Democratic Republic of the Congo" OR Djibouti OR "French Somaliland" OR Somalialand OR Somalia OR "Equatorial Guinea" OR Eritrea OR Ethiopia OR Gabon OR "Gabonese Republic" OR Gambia OR "The Gambia" OR Ghana OR "Gold Coast" OR Guinea OR Guinea-Bissau OR Kenya OR Lesotho OR Basutoland OR Liberia OR Madagascar OR "Malagasy Republic" OR Malawi OR Nyasaland OR Mali OR Mauritania OR Mauritius OR "Agalega Islands" OR Mozambique OR Namibia OR Niger OR Nigeria OR Rwanda OR Ruanda OR "South Africa" OR "Sao Tome" OR "Sierra Leone" OR Somalia OR Sudan OR Swaziland OR Eswatini OR Senegal OR Sudan OR "South Sudan" OR Seychelles OR Principe OR Tanzania OR Togo OR "Togolese Republic" OR Uganda OR Zambia OR Zimbabwe OR Rhodesia).mp. | 577110 |
| **4** | 1 and 2 and 3 | 427 |

*[mp=title, abstract, original title, name of substance word, subject heading word, floating sub-heading word, keyword heading word, organism supplementary concept word, protocol supplementary concept word, rare disease supplementary concept word, unique identifier, synonyms]*

**Embase 1974 to present**

| **#** | **Search terms** | **Results** |
| --- | --- | --- |
| **1** | ("referral" OR "patient pathway" OR "patient-pathway" OR "clinical pathway" OR "care continuum" OR "continuum of care" OR "continuum of patient care" OR "care continuity" OR "continuity of care" OR "continuity of patient care" OR "patient journey" OR "treatment cascade" OR "care cascade" OR "case management" OR "care management" OR "care pathway" OR "treatment pathway" OR "care delivery" OR "care network" OR "link to care" OR "linkage to care" OR "links to care" OR "health seeking" OR "health access" OR "healthcare access" OR "health care access" OR "medical access" OR "care seeking" OR "care access" OR "access to care" OR "access to healthcare" OR "access to health care").mp. | 676420 |
| **2** | ("neglected tropical disease" OR "NTD" OR "schisto*" OR "bilharzia" OR "snail fever" OR "mansoni" OR "japonicum" OR "mekongi" OR "guineensis" OR "s. intercalatum" OR "haematobium" OR "leprosy" OR "hansen’s disease" OR "buruli ulcer" OR "snakebite" OR "mycobacterium ulcerans" OR "chagas" OR "trypanosome cruzi" OR "american trypanosomiasis" OR "dengue" OR "chikungunya" OR "dracuncul*" OR "guinea-worm" OR "guineaworm" OR "echinococc*" OR "foodborne trematodiases" OR "foodborne trematode infection" OR "clonorchiasis" OR "opisthorchiasis" OR "fascioliasis" OR "paragonimiasis" OR "human african trypanosomiasis" OR "sleeping sickness" OR "HAT" OR "trypanosome" OR "leishmaniasis" OR "kala azar" OR "lymphatic filariasis" OR "elephantiasis" OR "podoconisis" OR "wuchereria bancrofti" OR "brugia" OR "onchocerc*" OR "river blindness" OR "rabies" OR "scabies" OR "soil transmitted helminthiasis" OR "roundworm" OR "ascaris" OR "whipworm" OR "trichuris" OR "hookworm" OR "necator americanus" OR "ancylostoma duodenale" OR "taenia*" OR "cysticercosis" OR "tapeworm" OR "mycetoma" OR "chromoblastomycosis" OR "deep mycoses" OR "trachoma" OR "endemic treponematoses" OR "treponem*" OR "yaws" OR "framboesia" OR "endemic syphilis" OR "bejel" OR "pinta").mp. | 328611 |
| **3** | (Africa OR "Sub-Saharan Africa" OR Angola OR Benin OR Botswana OR "Burkina Faso" OR "Burkina Fasso" OR "Burkina Faso" OR "Upper Volta" OR Burundi OR Urundi OR Cameroon OR Cameroons OR Cameron OR Camerons OR "Cape Verde" OR "Cabo Verde" OR "Central African Republic" OR Chad OR Comoros OR "Comoro Islands" OR Comores OR Congo OR Zaire OR "Cote d'Ivoire" OR "Ivory Coast" OR "DRC" OR "Democratic Republic of the Congo" OR Djibouti OR "French Somaliland" OR Somalialand OR Somalia OR "Equatorial Guinea" OR Eritrea OR Ethiopia OR Gabon OR "Gabonese Republic" OR Gambia OR "The Gambia" OR Ghana OR "Gold Coast" OR Guinea OR Guinea-Bissau OR Kenya OR Lesotho OR Basutoland OR Liberia OR Madagascar OR "Malagasy Republic" OR Malawi OR Nyasaland OR Mali OR Mauritania OR Mauritius OR "Agalega Islands" OR Mozambique OR Namibia OR Niger OR Nigeria OR Rwanda OR Ruanda OR "South Africa" OR "Sao Tome" OR "Sierra Leone" OR Somalia OR Sudan OR Swaziland OR Eswatini OR Senegal OR Sudan OR "South Sudan" OR Seychelles OR Principe OR Tanzania OR Togo OR "Togolese Republic" OR Uganda OR Zambia OR Zimbabwe OR Rhodesia).mp. | 604719 |
| **4** | 1 and 2 and 3 | 1078 |

*[mp=title, abstract, original title, name of substance word, subject heading word, floating sub-heading word, keyword heading word, organism supplementary concept word, protocol supplementary concept word, rare disease supplementary concept word, unique identifier, synonyms]*

**Global Health 1973 to present**

| **#** | **Search terms** | **Results** |
| --- | --- | --- |
| **1** | ("referral" OR "patient pathway" OR "patient-pathway" OR "clinical pathway" OR "care continuum" OR "continuum of care" OR "continuum of patient care" OR "care continuity" OR "continuity of care" OR "continuity of patient care" OR "patient journey" OR "treatment cascade" OR "care cascade" OR "case management" OR "care management" OR "care pathway" OR "treatment pathway" OR "care delivery" OR "care network" OR "link to care" OR "linkage to care" OR "links to care" OR "health seeking" OR "health access" OR "healthcare access" OR "health care access" OR "medical access" OR "care seeking" OR "care access" OR "access to care" OR "access to healthcare" OR "access to health care").mp. | 47118 |
| **2** | ("neglected tropical disease" OR "NTD" OR "schisto*" OR "bilharzia" OR "snail fever" OR "mansoni" OR "japonicum" OR "mekongi" OR "guineensis" OR "s. intercalatum" OR "haematobium" OR "leprosy" OR "hansen’s disease" OR "buruli ulcer" OR "snakebite" OR "mycobacterium ulcerans" OR "chagas" OR "trypanosome cruzi" OR "american trypanosomiasis" OR "dengue" OR "chikungunya" OR "dracuncul*" OR "guinea-worm" OR "guineaworm" OR "echinococc*" OR "foodborne trematodiases" OR "foodborne trematode infection" OR "clonorchiasis" OR "opisthorchiasis" OR "fascioliasis" OR "paragonimiasis" OR "human african trypanosomiasis" OR "sleeping sickness" OR "HAT" OR "trypanosome" OR "leishmaniasis" OR "kala azar" OR "lymphatic filariasis" OR "elephantiasis" OR "podoconisis" OR "wuchereria bancrofti" OR "brugia" OR "onchocerc*" OR "river blindness" OR "rabies" OR "scabies" OR "soil transmitted helminthiasis" OR "roundworm" OR "ascaris" OR "whipworm" OR "trichuris" OR "hookworm" OR "necator americanus" OR "ancylostoma duodenale" OR "taenia*" OR "cysticercosis" OR "tapeworm" OR "mycetoma" OR "chromoblastomycosis" OR "deep mycoses" OR "trachoma" OR "endemic treponematoses" OR "treponem*" OR "yaws" OR "framboesia" OR "endemic syphilis" OR "bejel" OR "pinta").mp. | 213480 |
| **3** | (Africa OR "Sub-Saharan Africa" OR Angola OR Benin OR Botswana OR "Burkina Faso" OR "Burkina Fasso" OR "Burkina Faso" OR "Upper Volta" OR Burundi OR Urundi OR Cameroon OR Cameroons OR Cameron OR Camerons OR "Cape Verde" OR "Cabo Verde" OR "Central African Republic" OR Chad OR Comoros OR "Comoro Islands" OR Comores OR Congo OR Zaire OR "Cote d'Ivoire" OR "Ivory Coast" OR "DRC" OR "Democratic Republic of the Congo" OR Djibouti OR "French Somaliland" OR Somalialand OR Somalia OR "Equatorial Guinea" OR Eritrea OR Ethiopia OR Gabon OR "Gabonese Republic" OR Gambia OR "The Gambia" OR Ghana OR "Gold Coast" OR Guinea OR Guinea-Bissau OR Kenya OR Lesotho OR Basutoland OR Liberia OR Madagascar OR "Malagasy Republic" OR Malawi OR Nyasaland OR Mali OR Mauritania OR Mauritius OR "Agalega Islands" OR Mozambique OR Namibia OR Niger OR Nigeria OR Rwanda OR Ruanda OR "South Africa" OR "Sao Tome" OR "Sierra Leone" OR Somalia OR Sudan OR Swaziland OR Eswatini OR Senegal OR Sudan OR "South Sudan" OR Seychelles OR Principe OR Tanzania OR Togo OR "Togolese Republic" OR Uganda OR Zambia OR Zimbabwe OR Rhodesia).mp. | 307069 |
| **4** | 1 and 2 and 3 | 395 |

*[mp=title, abstract, original title, name of substance word, subject heading word, floating sub-heading word, keyword heading word, organism supplementary concept word, protocol supplementary concept word, rare disease supplementary concept word, unique identifier, synonyms]*

**Global Index Medicus 1901 to present**

| **#** | **Search terms** | **Results** |
| --- | --- | --- |
| **1** | tw: ("referral" or "patient pathway" or "patient-pathway" or "clinical pathway" or "care continuum" or "continuum of care" or "continuum of patient care" or "care continuity" or "continuity of care" or "continuity of patient care" or "patient journey" or "treatment cascade" or "care cascade" or "case management" or "care management" or "care pathway" or "treatment pathway" or "care delivery" or "care network" or "link to care" or "linkage to care" or "links to care" or "health seeking" or "health access" or "healthcare access" or "health care access" or "medical access" or "care seeking" or "care access" or "access to care" or "access to healthcare" or "access to health care") | 41413 |
| **2** | tw: ("neglected tropical disease" or "NTD" or "schisto*" or "bilharzia" or "snail fever" or "mansoni" or "japonicum" or "mekongi" or "guineensis" or "s. intercalatum" or "haematobium" or "leprosy" or "hansen’s disease" or "buruli ulcer" or "snakebite" or "mycobacterium ulcerans" or "chagas" or "trypanosome cruzi" or "american trypanosomiasis" or "dengue" or "chikungunya" or "dracuncul*" or "guinea-worm" or "guineaworm" or "echinococc*" or "foodborne trematodiases" or "foodborne trematode infection" or "clonorchiasis" or "opisthorchiasis" or "fascioliasis" or "paragonimiasis" or "human african trypanosomiasis" or "sleeping sickness" or "HAT" or "trypanosome" or "leishmaniasis" or "kala azar" or "lymphatic filariasis" or "elephantiasis" or "podoconisis" or "wuchereria bancrofti" or "brugia" or "onchocerc*" or "river blindness" or "rabies" or "scabies" or "soil transmitted helminthiasis" or "roundworm" or "ascaris" or "whipworm" or "trichuris" or "hookworm" or "necator americanus" or "ancylostoma duodenale" or "taenia*" or "cysticercosis" or "tapeworm" or "mycetoma" or "chromoblastomycosis" or "deep mycoses" or "trachoma" or "endemic treponematoses" or "treponem*" or "yaws" or "framboesia" or "endemic syphilis" or "bejel" or "pinta") | 52805 |
| **3** | tw: (Africa or "Sub-Saharan Africa" or Angola or Benin or Botswana or "Burkina Faso" or "Burkina Fasso" or "Burkina Faso" or "Upper Volta" or Burundi or Urundi or Cameroon or Cameroons or Cameron or Camerons or "Cape Verde" or "Cabo Verde" or "Central African Republic" or Chad or Comoros or "Comoro Islands" or Comores or Congo or Zaire or "Cote d'Ivoire" or "Ivory Coast" or "DRC" or "Democratic Republic of the Congo" or Djibouti or "French Somaliland" or Somalialand or Somalia or "Equatorial Guinea" or Eritrea or Ethiopia or Gabon or "Gabonese Republic" or Gambia or "The Gambia" or Ghana or "Gold Coast" or Guinea or Guinea-Bissau or Kenya or Lesotho or Basutoland or Liberia or Madagascar or "Malagasy Republic" or Malawi or Nyasaland or Mali or Mauritania or Mauritius or "Agalega Islands" or Mozambique or Namibia or Niger or Nigeria or Rwanda or Ruanda or "South Africa" or "Sao Tome" or "Sierra Leone" or Somalia or Sudan or Swaziland or Eswatini or Senegal or Sudan or "South Sudan" or Seychelles or Principe or Tanzania or Togo or "Togolese Republic" or Uganda or Zambia or Zimbabwe or Rhodesia) | 40717 |
| **4** | 1 and 2 and 3 | 22 |

*[tw=title,abstract,subject]*

**Web of Science – Core Collection, Science Citation Index Expanded 1900 to present**

| **#** | **Search terms** | **Results** |
| --- | --- | --- |
| **1** | AB=("referral" or "patient pathway" or "patient-pathway" or "clinical pathway" or "care continuum" or "continuum of care" or "continuum of patient care" or "care continuity" or "continuity of care" or "continuity of patient care" or "patient journey" or "treatment cascade" or "care cascade" or "case management" or "care management" or "care pathway" or "treatment pathway" or "care delivery" or "care network" or "link to care" or "linkage to care" or "links to care" or "health seeking" or "health access" or "healthcare access" or "health care access" or "medical access" or "care seeking" or "care access" or "access to care" or "access to healthcare" or "access to health care")) | 140557 |
| **2** | AB=("neglected tropical disease" or "NTD" or "schisto*" or "bilharzia" or "snail fever" or "mansoni" or "japonicum" or "mekongi" or "guineensis" or "s. intercalatum" or "haematobium" or "leprosy" or "hansen’s disease" or "buruli ulcer" or "snakebite" or "mycobacterium ulcerans" or "chagas" or "trypanosome cruzi" or "american trypanosomiasis" or "dengue" or "chikungunya" or "dracuncul*" or "guinea-worm" or "guineaworm" or "echinococc*" or "foodborne trematodiases" or "foodborne trematode infection" or "clonorchiasis" or "opisthorchiasis" or "fascioliasis" or "paragonimiasis" or "human african trypanosomiasis" or "sleeping sickness" or "HAT" or "trypanosome" or "leishmaniasis" or "kala azar" or "lymphatic filariasis" or "elephantiasis" or "podoconisis" or "wuchereria bancrofti" or "brugia" or "onchocerc*" or "river blindness" or "rabies" or "scabies" or "soil transmitted helminthiasis" or "roundworm" or "ascaris" or "whipworm" or "trichuris" or "hookworm" or "necator americanus" or "ancylostoma duodenale" or "taenia*" or "cysticercosis" or "tapeworm" or "mycetoma" or "chromoblastomycosis" or "deep mycoses" or "trachoma" or "endemic treponematoses" or "treponem*" or "yaws" or "framboesia" or "endemic syphilis" or "bejel" or "pinta") | 148,317 |
| **3** | AB=(Africa or "Sub-Saharan Africa" or Angola or Benin or Botswana or "Burkina Faso" or "Burkina Fasso" or "Burkina Faso" or "Upper Volta" or Burundi or Urundi or Cameroon or Cameroons or Cameron or Camerons or "Cape Verde" or "Cabo Verde" or "Central African Republic" or Chad or Comoros or "Comoro Islands" or Comores or Congo or Zaire or "Cote d'Ivoire" or "Ivory Coast" or "DRC" or "Democratic Republic of the Congo" or Djibouti or "French Somaliland" or Somalialand or Somalia or "Equatorial Guinea" or Eritrea or Ethiopia or Gabon or "Gabonese Republic" or Gambia or "The Gambia" or Ghana or "Gold Coast" or Guinea or Guinea-Bissau or Kenya or Lesotho or Basutoland or Liberia or Madagascar or "Malagasy Republic" or Malawi or Nyasaland or Mali or Mauritania or Mauritius or "Agalega Islands" or Mozambique or Namibia or Niger or Nigeria or Rwanda or Ruanda or "South Africa" or "Sao Tome" or "Sierra Leone" or Somalia or Sudan or Swaziland or Eswatini or Senegal or Sudan or "South Sudan" or Seychelles or Principe or Tanzania or Togo or "Togolese Republic" or Uganda or Zambia or Zimbabwe or Rhodesia) | 389053 |
| **4** | 1 and 2 and 3 | 223 |

*[AB=abstract]*

**Cochrane Central Register of Controlled Trials (CENTRAL) 1996 to present**

| **#** | **Search terms** | **Results** |
| --- | --- | --- |
| **1** | ("referral" or "patient pathway" or "patient-pathway" or "clinical pathway" or "care continuum" or "continuum of care" or "continuum of patient care" or "care continuity" or "continuity of care" or "continuity of patient care" or "patient journey" or "treatment cascade" or "care cascade" or "case management" or "care management" or "care pathway" or "treatment pathway" or "care delivery" or "care network" or "link to care" or "linkage to care" or "links to care" or "health seeking" or "health access" or "healthcare access" or "health care access" or "medical access" or "care seeking" or "care access" or "access to care" or "access to healthcare" or "access to health care"):ti,ab,kw | 29223 |
| **2** | ("neglected tropical disease" or "NTD" or "schisto*" or "bilharzia" or "snail fever" or "mansoni" or "japonicum" or "mekongi" or "guineensis" or "s. intercalatum" or "haematobium" or "leprosy" or "hansen’s disease" or "buruli ulcer" or "snakebite" or "mycobacterium ulcerans" or "chagas" or "trypanosome cruzi" or "american trypanosomiasis" or "dengue" or "chikungunya" or "dracuncul*" or "guinea-worm" or "guineaworm" or "echinococc*" or "foodborne trematodiases" or "foodborne trematode infection" or "clonorchiasis" or "opisthorchiasis" or "fascioliasis" or "paragonimiasis" or "human african trypanosomiasis" or "sleeping sickness" or "HAT" or "trypanosome" or "leishmaniasis" or "kala azar" or "lymphatic filariasis" or "elephantiasis" or "podoconisis" or "wuchereria bancrofti" or "brugia" or "onchocerc*" or "river blindness" or "rabies" or "scabies" or "soil transmitted helminthiasis" or "roundworm" or "ascaris" or "whipworm" or "trichuris" or "hookworm" or "necator americanus" or "ancylostoma duodenale" or "taenia*" or "cysticercosis" or "tapeworm" or "mycetoma" or "chromoblastomycosis" or "deep mycoses" or "trachoma" or "endemic treponematoses" or "treponem*" or "yaws" or "framboesia" or "endemic syphilis" or "bejel" or "pinta"):ti,ab,kw | 7340 |
| **3** | (Africa or "Sub-Saharan Africa" or Angola or Benin or Botswana or "Burkina Faso" or "Burkina Fasso" or "Burkina Faso" or "Upper Volta" or Burundi or Urundi or Cameroon or Cameroons or Cameron or Camerons or "Cape Verde" or "Cabo Verde" or "Central African Republic" or Chad or Comoros or "Comoro Islands" or Comores or Congo or Zaire or "Cote d'Ivoire" or "Ivory Coast" or "DRC" or "Democratic Republic of the Congo" or Djibouti or "French Somaliland" or Somalialand or Somalia or "Equatorial Guinea" or Eritrea or Ethiopia or Gabon or "Gabonese Republic" or Gambia or "The Gambia" or Ghana or "Gold Coast" or Guinea or Guinea-Bissau or Kenya or Lesotho or Basutoland or Liberia or Madagascar or "Malagasy Republic" or Malawi or Nyasaland or Mali or Mauritania or Mauritius or "Agalega Islands" or Mozambique or Namibia or Niger or Nigeria or Rwanda or Ruanda or "South Africa" or "Sao Tome" or "Sierra Leone" or Somalia or Sudan or Swaziland or Eswatini or Senegal or Sudan or "South Sudan" or Seychelles or Principe or Tanzania or Togo or "Togolese Republic" or Uganda or Zambia or Zimbabwe or Rhodesia):ti,ab,kw | 31857 |
| **4** | 1 and 2 and 3 | 33 |

*[ti,ab,kw=title, abstract, keywords were searched for terms]*
