## Supplementary material for "Conceptualising care pathways for neglected tropical diseases in sub-Saharan Africa: A systematic scoping review": S3 Data dictionary

**S3 Text: Data Dictionary**

|  | **Variable** | **Definition** | **Proposed Options** |
| --- | --- | --- | --- |
| **1** | **title** | Study title | [Title] |
| **2** | **author_year** | Last name of first author | [Author name] |
| **3** | **publication_year** | Year the study was published | [Year] |
| **4** | **study_year** | Year study was conducted (or began if study duration >1 year) | [Year] |
| **5** | **study_duration** | How long was the study (in months)? | [Years] |
| **6** | **country** | In which country did the study take place? | Sub-Saharan Africa |
| **7** | **country_region** | In what region within the study country? | [Describe] |
| **8** | **locality** | In what locality did the study take place? | Rural, Peri-Urban, Urban, Mixed |
| **9** | **study_setting** | What was the study setting? | Community, Facility, Mixed |
| **10** | **study_aims** | What were the research aims(s) of the study? | [Describe] |
|  | **study_aim_categorisation** | [Not extracted – grouping of study aims by type post data extraction] | Biomedical research providing diagnosis and treatment (patient care) to study participants, Patient experience(s) of diagnosis, treatment, and care continuity, Routine in-facility case management, including case studies |
| **11** | **study_design** | What was the design of the study? | Prospective cohort, retrospective cohort, case study, case-control, before and after study, cross-sectional, comparative cross-sectional, repeat cross-sectional, Randomized Controlled Trial |
| **12** | **study_data_collection** | By what methodology was the data collected? | Survey, interviews, focus groups, clinical observation |
| **13** | **study_design_data_type** | Was the study collecting qualitative, quantitative, or mixed methodology data? | Quantitative, qualitative, mixed methodology |
| **14** | **study_intervention** | If the study was a randomised controlled trial or involved an evaluation of an intervention, what was the intervention? | [Describe] |
| **15** | **study_comparison** | If the study included an intervention what was the control group/comparison? | [Describe] |
|  | **Study_ntd_category** | [Not extracted – grouping of NTDs by type post data extraction] | PC-NTD, morbidity management, Snakebite/Animal bite injury |
| **16** | **study_disease_main** | What was the primary disease being investigated in the study? | All NTDs |
| **17** | **study_disease_secondary** | What was the secondary disease being investigated in the study? | All NTDs |
| **18** | **study_disease_other** | Were other NTD(s) being investigated in the study? | All NTDs |
| **19** | **study_disease_other_2** | Were other NTD(s) being investigated in the study? | All NTDs |
| **20** | **study_condition_non_NTD** | Was the disease being managed alongside any other health conditions/co-morbidites (not NTDs)? | All diseases and health conditions, (excluding NTDs) |
| **21** | **study_symptoms** | For studies focused on symptoms of multiple aetiologies, what symptoms were investigated in the study? | Anaemia, blindness, skin ulcers, fever, diarrhoea, etc |
| **22** | **sample_size** | What is the number participants in study sample? | Total number of participants |
| **23** | **age_groups** | What were the age groups of the study participants? | Children, Adults, Children and Adults/All |
| **24** | **study_participant_patients** | Were current (suspect or confirmed) or former patients participants in the study? Participants in a screening campaign/prevalence study count as suspect patients. | Suspect patients, Suspect patients (screening campaign), Current patients, former patients |
| **25** | **study_participants_other** | Aside from patients, who were the other participants in the study? | Family members of current or former patients, Community Health Workers, Clinicians, Key Informants (NGO workers, Ministry of Health Staff), etc |
| **26** | **chws_yn** | Were Community Health Workers explicitly involved in the study? | Yes, No |
| **27** | **chws_titles** | If Community Health Workers were involved in the study, what were they referred to as? | Community Health Workers, Community Health Extension Workers, Community Medicine Distributors, Community Health Assistants, Community Health Promoters, Community Medicine Distributors, Village Health Team members, etc |
| **28** | **chws_definitions** | If Community Health Workers were invovled in the study, how were they defined or identified? | [Definition of CHWs], Unspecified |
| **29** | **chws_roles_research_yn** | If Community Health Workers were involved in the study, did they play a role in research facilitation? | Yes, No, Unspecified, N/A |
| **30** | **chws_roles_ research** | If Community Health Workers were involved in the study and played a role in research facilitation, what was that role? | Study outreach, recruitment of study participants, data collection, etc. |
| **31** | **chws_compensation** | If Community Health Workers were involved in the study and played a role in research facilitation, did the study describe how they were compensated? Explain. | Yes, No, Unspecified, N/A |
| **32** | **chws_roles_care_yn** | If Community Health Workers were involved in the study, did they play a role in patient care? | Yes, No, Unspecified, N/A |
| **33** | **chws_roles_care** | If Community Health Workers were involved in the study and played a role in patient care, what was that role? | Delivery of medications, follow up on referrals, participation in health promotion, etc |
| **34** | **chws_training** | If Community Health Workers were involved in the study, describe any training they received. | [Describe] |
| **35** | **patient_pathways_yn** | Did the study explicitly describe a sequence of events related to diagnosis, treatment, and follow up from the patient perspective (the patient pathway)? | [Describe] |
| **36** | **patient_pathways_terminology** | If "yes" to variable patient_pathways_yn, what was the terminology used to describe the sequence of events related to diagnosis, treatment, and follow up from the patient perspective (the patient pathway)? | [Describe] |
| **37** | **patient_pathways_depiction** | If "yes" to variable patient_pathways_yn, how as the patient pathway depicted? | [Describe] |
| **38** | **patient_pathways_start** | If "yes" to variable patient_pathways_yn, what was considered the start of the pathway? | [Describe] |
| **39** | **patient_pathways_end** | If "yes" to variable patient_pathways_yn, what was considered the end of the pathway? | [Describe] |
| **40** | **patient_pathways_single** | If "yes" to variable patient_pathways_yn, was there one primary patient pathway presented? | Yes, No |
| **41** | **patient_pathways_multiple** | If "yes" to variable patient_pathways_yn, were multiple possible patient pathways presented and/or were the pathways non-linear (i.e. multiple attempts seeking diagnostics from different providers prior to treatment)? | Yes, No |
| **42** | **patient_pathway_notes** | **For all studies,** where any other details regarding the patient pathway and/or healthcare seeking in health system? | [Describe] |
| **43** | **diagnosis_misdiagnosis** | Did the study explicitly reference misdiagnosis, treatment for another disease, or discordant results? | Yes, No |
| **44** | **diagnosis_notes** | Any other information regarding the diagnosis/suspect case assessment? | [Describe] |
| **45** | **treatment_ notes** | Any other information regarding the treatment, including any additional levels of treatment or why specific treatment options were chosen? | [Explain] |
| **46** | **referrals_yn** | Did the study explicitly mention referrals? | No, Yes, Yes but only as part of ethics statement |
| **47** | **referrals_why** | If the study explicitly mentioned referrals, why was the referral was given? i.e. Why the referral was necessary or why care could not be provided locally | Treatment following prevalence study, Treatment not available locally or immediately, Suspect case in need of diagnostic confirmation |
| **48** | **referrals_where** | If the study explicitly mentioned referrals, what was the physical setting where the referral was given? | Community (communal location), community (home), school, primary, secondary, or tertiary health facility, other |
| **49** | **referrals_who_provided** | If the study explicitly mentioned referrals, who provided the referral? | Community health worker, nurse, primary care physician, case manager (non-clinical) |
| **50** | **referrals_other_actors** | If the study explicitly mentioned referrals, were there other actors who played a role in case management or coordination of the referral? | Commmunity Health Workers, family members, nurses, case manager, medical doctor |
| **51** | **referrals_to** | Where was the referral sending the patient for follow up care? | Primary, secondary, tertiary; 1) HC II (Parish-level), 2) HC III (Sub-County-level), 3) HC IV (County-level), 4) General Hospital (GH), 5) Regional Referral Hospital (RRH), and 6) National Referral Hospital (NRH), Hospital (tier unspecified), pharmacy, other/unspecified |
| **52** | **referral_success** | Was any information provided regarding the success of the referral? | Yes [Explain], No, N/A, Unspecified |
| **53** | **referral_notes** | Any other notes regarding the referral? | [Describe] |
| **54** | **care_continuity** | Does the study explicitly mention a strategy for follow up for study participants? If yes, what was that strategy? | Case management support, transportation, financial support/free care provision, etc. |
| **55** | **care_continuity_enablers** | Did the study describe any other factors that enabled access to and/or continuity of care? If Yes, describe. | [Describe] |
| **56** | **care_continuity_inhibitors** | Did the study describe any other factors that inhibited access to and/or continuity of care? IF yes, describe. | [Describe] |
| **57** | **care_continuity_recommendations** | Did the study propose any solutions or recommendations for identified barriers to care? | [i.e. Transportation support, dedicated case management, capacity building of clinicians, construction of additional health facilities] |
| **58** | **ethical_challenges_yn** | Did the study explicitly mention any ethical challenges in the study? | Yes, No |
| **59** | **ethical_challenges** | If the study explicitly mentioned any ethical challenges, what were they and how were they handled? | [Describe] |
| **60** | **study_other** | Any other notes regarding the study? | [Describe] |
