## Supplementary material for "Conceptualising care pathways for neglected tropical diseases in sub-Saharan Africa: A systematic scoping review": S4 References for included studies

**S4_References included**

60. [Godwin-Akpan T.G., Chowdhury S., Rogers E.J., Kollie K.K., Zaizay F.Z., Wickenden A., et al. Recommendations for an Optimal Model of integrated case detection, referral, and confirmation of Neglected Tropical Diseases: A case study in Bong County, Liberia. medRxiv [Internet]. 2022;((Godwin-Akpan, Wickenden) AIM Initiative/American Leprosy Mission, Greenville, SC, United States(Chowdhury, Dean) Department of International Public Health, Liverpool School of Tropical Medicine, Liverpool, United Kingdom(Rogers, Kollie, Zaizay) Neglected). Available from: https://www.medrxiv.org/](https://www.medrxiv.org/)

77. Koka E., Abdulai H.B.T. Health seeking behaviour for buruli ulcer disease in the OBOM sub-district of the GA south municipality of Ghana. American Journal of Tropical Medicine and Hygiene. 2020;103(5 SUPPL):10.

148. Tora A., Davey G., Tadele G. Factors related to discontinued clinic attendance by patients with podoconiosis in southern Ethiopia: a qualitative study. BMC public health. 2012;12((Tora) Department of Sociology, Wolaita Sodo University, Sodo, Ethiopia.):902.
